# Public perceptions and preventive behaviours during the early phase of the COVID-19 pandemic: a comparative study between Hong Kong and the United Kingdom

**DOI:** 10.1101/2020.08.06.20169409

**Authors:** LR Bowman, KO Kwok, RE Redd, YY Yi, H Ward, WI Wei, C Atchison, SYS Wong

**Author notes:** Authors contributed equally (LR Bowman, KO Kwok). Authors contributed equally (C Atchison, SYS Wong). Joint correspondence.

## Abstract

**Background:** In the absence of treatments and vaccines, the mitigation of COVID-19 relies on population engagement in non-pharmaceutical interventions, which is driven by their risk perception, anxiety level and knowledge. There may also be regional discrepancies in these drivers due to different historical exposure to disease outbreaks, government responses and cultures. As such, this study compared psycho-behavioral responses in two regions during the early phase of the pandemic.

**Methods:** Comparable cross-sectional surveys were administered among adults in Hong Kong (HK) and the United Kingdom (UK) during the early phase of each respective epidemic. Explanatory variables included demographics, risk perception and knowledge of COVID-19, anxiety level and preventive behaviors. Responses were weighted according to census data. Logistic regression models, including interaction terms to quantify regional differences, were used to assess the association between explanatory variables and the adoption of social-distancing measures.

**Results:** Data of 3431 complete responses (HK:1663; UK:1768) were analysed. Perceived severity differed by region (HK: 97.5%; UK: 20.7%). A large proportion of respondents were abnormally/borderline anxious (HK:64.8%; UK:45.9%) and regarded direct contact with infected individuals as the transmission route of COVID-19 (HK:94.0-98.5%; UK:69.2-93.5%), with HK identifying additional routes. HK reported high levels of adoption of social-distancing (HK:32.4-93.7%; UK:17.6-59.0%) and mask-wearing (HK:98.8%; UK:3.1%). The impact of perceived severity and perceived ease of transmission on the adoption of social-distancing varied by region. In HK, they had no impact, whereas in the UK, those who perceived severity as “high” were more likely to adopt social-distancing (aOR:1.58-3.01), and those who perceived transmission as “easy” were prone to both general social-distancing (aOR:2.00, 95% CI:1.57, 2.55) and contact avoidance (aOR:1.80, 95% CI: 1.41, 2.30). The impact of anxiety on adopting social-distancing did not vary by region.

**Discussion:** These results suggest that health officials should ascertain and consider baseline levels of risk perception and knowledge in the populations, as well as prior sensitisation to infectious disease outbreaks, during the development of mitigation strategies. Risk communication should be done through suitable media channels - and trust should be maintained - while early intervention remains the cornerstone of effective outbreak response.

## INTRODUCTION

In December 2019, a novel coronavirus (SARS-CoV-2) emerged in Wuhan, Hubei province,

China, and spread rapidly throughout countries worldwide, forming the second pandemic of the 21^st^ century **[1]**. The disease progression varies by regions. As of 2 August 2020, there have been at least 17 million cases and over 670,000 deaths worldwide **[2]**.

Prior to the availability of effective treatments and vaccines, strategies to mitigate the impact of the pandemic have been primarily non-pharmaceutical **[3]**, mainly focused on public health promotion of the use of simple but effective preventive measures **[4, 5]**. Many important control strategies currently promoted by governments need public participation, either through direct adoption of preventative behaviours, such as handwashing or wearing face masks, or through compliance with social distancing policies, such as recommendations to avoid public transport and mass gatherings.

Previous studies of severe acute respiratory syndrome (SARS) and pandemic influenza have shown that governments should account for risk perception and anxiety when promoting preventative measures. There is evidence that higher perceived risk of infection is associated with increased adoption of precautionary measures against infection **[6, 7]**, while increased anxiety has also been shown to increase the likelihood that people will engage in protective behaviour **[8]**. Moreover, longitudinal data suggest that such perceptions, behaviours and anxieties change with context, and over time, as uncertainty about disease severity reduces, and knowledge of transmission increases **[9]**.

During the current coronavirus disease 2019 (COVID-19) pandemic, several studies have examined public risk perceptions and knowledge in various countries, including Finland **[10]**, Israel **[11]**, Italy **[12]**, Nigeria **[13]**, the United States **[14]** and Vietnam **[15]**, but only few have identified the factors associated with greater adoption of preventative measures, or how these associations vary by context. In HK, both greater understanding of COVID-19 and increased anxiety were associated with greater adoption of social distancing behaviours **[5]**; whereas in the UK, there was a significant socio-economic gradient in the ability to adopt and comply with social distancing measures, specifically the ability to work from home and ability to self-isolate **[16]**.

This initial evidence - that there is variation across context in affective responses, risk perceptions, the impact of socio-demographic factors, and their association with uptake of preventative behaviours - has significant implications when tailoring policy to specific circumstances. In order to tease out these relationships, a more thorough, comparative analysis is required. Yet studies in different countries often use slightly different metrics to measure the same behaviour, which can lead to difficulty when interpreting the significance of heterogeneous contexts.

In this study, we examined and compared public perception and adoption of preventive behaviours in response to the early phase of the COVID-19 pandemic in two different regions: HK and the UK. We further investigated the factors associated with greater adoption of different types of social distancing measures. Our results have immediate implications on how health officials plan and communicate the mitigation strategies for the ongoing COVID-19 pandemic to communities.

## MATERIALS AND METHODS

### Study design and recruitment

In HK and the UK, cross-sectional surveys were conducted during the early phase of the COVID-19 pandemic, when there were only limited government-level interventions in place **[5, 16]**. The survey period in HK was from 24 January 2020 to 13 February 2020, and in the UK from 17 to 18 March 2020. In HK, the first laboratory-confirmed case was reported on 23 January 2020, rising to 53 cases by 13 February 2020 **[17]**; whereas in the UK, the first two laboratory-confirmed cases were reported on 30 January 2020, rising to 3,902 cases by 17 March 2020 **[18]**. In HK, an online survey was distributed by district councils to their residents. Those individuals aged 18 years or above, who understood Chinese and lived in HK were eligible to participate **[5]**. In the UK, the online survey was distributed by a market research company to a random sample of an existing panel of 800,000+ adults (aged ≥ 18 years) **[16]**. Details of the survey design and sampling strategy are described elsewhere **[5, 16]**.

### Study instrument

The questionnaire used was largely similar across the two regions. Sociodemographic variables included age, sex, educational attainment and employment status. Anxiety level was measured using the Hospital, Anxiety and Depression scale - Anxiety (HASD-A) (0-7 = Normal; 8-10 = Borderline abnormal; 11-21 = Abnormal) **[19]**. Risk perception towards COVID-19 was measured by perceived severity of symptoms if infected with COVID-19. Knowledge of COVID-19 was assessed by asking whether COVID-19 could be transmitted through various routes, including direct human exposure (for example, physical contact or a face-to-face conversation with someone who has coronavirus with or without symptoms) and other types of exposure (for example, visiting wet markets, or consumption of wild animal meat). Respondents were also asked about the sources from which they retrieved information about COVID-19. In addition, they were asked about adoption of preventative behaviours to cope with the transmission of COVID-19. Three types of preventative measures were considered: personal hygiene, social distancing and travel avoidance.

### Data analysis

Descriptive statistics for all variables present the number of respondents and the raw or weighted percentages. The responding samples were weighted to be representative of the UK (UK 2011 census data **[20]**) and HK adult population (HK 2016 census data **[21]**). Chi-square goodness-of-fit tests were used for comparison of characteristics across regions. Multivariate logistic regression models were used to identify sociodemographic and psychosocial factors associated with the adoption of three types of social distancing: (i) general measures, specified by avoiding crowded places, social events and going out; (ii) contact measures, specified by avoiding contact with individuals who had fever or respiratory symptoms, and who had been to affected areas recently; and (iii) work measure, specified by avoiding going to work.

Common and comparable sociodemographic factors considered in separate analytical studies **[5, 16]** were included in this comparative analysis. The association between psychosocial factors (including level of anxiety, perceived severity and perceived ease of transmission) and adoption of social-distancing measures was considered a priori to be affected by region. Therefore, we examined the effect modification due to region using interaction terms in the baseline models, which can be interpreted as the difference of the estimated effects of psychosocial factors on adopting social-distancing measures due to different regions. Adjusted odds ratios (aOR) and 95% confidence intervals (CI) were estimated. Associations with a p-value <0.05 in the adjusted analyses were considered to be statistically significant. Analyses were conducted in R (v3.6.3).

### Ethical Approval

The study was approved by Imperial College London Research Ethics Committee (reference number: 20IC5861) and Survey and Behavioral Research Ethics Committee of The Chinese University of Hong Kong (reference number: SBRE-19-625).

## RESULTS

### Demographic differences

There were significant differences in the sociodemographic characteristics of study respondents between the two regions. HK respondents were younger, with 26.0% aged 18-24 years, compared with 9.4% for the UK (*P*<0.001) (**Table 1**). The HK sample contained a greater proportion of females (68.6% [HK] vs. 52.9% [UK]; *P*<0.01), and those educated to university degree level or above (63.2% [HK] vs. 33.7% [UK]; *P*<0.001). Employment status reflected the age structure of respondents in each setting, with a greater proportion of the UK respondents in the retired category (2.6% [HK] vs. 27.2% [UK]; *P*<0.001) (**Table 1**).

**Table 1.** Hong Kong and United Kingdom study respondent characteristics

| Characteristics | UK (n = 1768) |  | HK (n = 1663) |  | P-value <sup>a</sup> |
| --- | --- | --- | --- | --- | --- |
|  | Number | % | Number | % |  |
| Age |  |  |  |  |  |
| 18-24 | 166 | 9.4 | 433 | 26.0 | <0.001 |
| 25-34 | 243 | 13.7 | 535 | 32.2 |  |
| 35-44 | 335 | 18.9 | 370 | 22.2 |  |
| 45-54 | 300 | 17.0 | 193 | 11.6 |  |
| 55+ | 724 | 41.0 | 132 | 7.9 |  |
| Sex |  |  |  |  |  |
| Female | 936 | 52.9 | 1141 | 68.6 | <0.001 |
| Male | 832 | 47.1 | 522 | 31.4 |  |
| Education attainment |  |  |  |  |  |
| No formal qualification/ lower secondary or below | 100 | 5.7 | 53 | 3.2 | <0.001 |
| Secondary level qualification/ higher secondary | 738 | 41.7 | 292 | 17.6 |  |
| Post-secondary but below degree | 334 | 18.9 | 267 | 16.1 |  |
| Degree or above | 596 | 33.7 | 1051 | 63.2 |  |
| Employment status |  |  |  |  |  |
| Employer/Employee | 1025 | 58.0 | 1135 | 68.3 | <0.001 |
| Full-time student | 90 | 5.1 | 278 | 16.7 |  |
| Unemployed/Not working | 172 | 9.7 | 206 | 12.4 |  |
| Retired | 481 | 27.2 | 44 | 2.6 |  |
| Perceived severity <sup>b</sup> |  |  |  |  |  |
| Level 1 | 96 | 5.4 | 1071 | 64.4 | <0.001 |
| Level 2 | 270 | 15.3 | 550 | 33.1 |  |
| Level 3 | 1058 | 59.8 | 32 | 1.9 |  |
| Level 4 | 320 | 18.1 | 7 | 0.4 |  |
| Level 5 | 24 | 1.4 | 3 | 0.2 |  |
| Worry about COVID-19 |  |  |  |  |  |
| Very worried | 536 | 30.3 | 852 | 51.2 | <0.001 |
| Fairly worried | 858 | 48.5 | 723 | 43.5 |  |
| Neutral/ Don't know | 5 | 0.3 | 40 | 2.4 |  |
| Not very worried | 295 | 16.7 | 1 | 0.1 |  |
| Not at all worried | 74 | 4.2 | 47 | 2.8 |  |
| Anxiety level |  |  |  |  |  |
| Normal | 956 | 54.1 | 586 | 35.2 | <0.001 |
| Borderline abnormal | 336 | 19.0 | 512 | 30.8 |  |
| Abnormal | 476 | 26.9 | 565 | 34.0 |  |

<sup>a</sup> Chi-square goodness-of-fit test
<sup>b</sup> Level 1 = very serious (HK) / life-threatening (UK); Level 2 = serious (HK) / severe (e.g. may need care and treatment in hospital) (UK); Level 3 = neutral (HK) / moderate (e.g. may need self-care and rest in bed) (UK); Level 4 = not serious (HK) / mild (e.g. can go about daily tasks normally) (UK); Level 5 = not serious at all (HK) / no symptoms (UK)

### Perceptions and beliefs

Higher perceived severity of COVID-19 was observed among HK respondents, with 97.5% rating the symptoms of COVID-19 infection as serious or very serious compared with only 20.7% of the UK respondents. In terms of levels of concern, 94.7% of the HK sample responded as feeling either very worried or fairly worried, compared with 78.8% of the UK sample. The HADS-A scores reflected similar trends, with 64.8% of the HK sample recording an abnormal or borderline abnormal result, compared with 45.9% of the UK sample (**Table 1**).

### Knowledge and information sources

The majority of respondents regarded direct contact with infected individuals or virus-contaminated environments as the primary means of virus transmission (HK: 94.0-98.5%; UK: 69.2-93.5%) (**Table 2**). However, respondents from HK identified a far boarder scope of transmission routes. A large proportion of HK respondents (66.5-93.3%) regarded wild animal meat, wet markets and imported goods as potential exposures, compared to 11.3-21.5% in the UK. There was also significant variation across use and reliability of information sources (**Table S1**)/The majority of respondents deemed health professionals reliable (HK, UK: >80%) but few could access them (HK:4.7%; UK:11.5%). In addition, most of the UK respondents (90.7%) considered official websites reliable, compared to 15.5% among HK respondents at the beginning of the pandemic.

**Table 2.** Knowledge of COVID-19 transmission

| Are the following transmission routes of COVID-19? | Number of respondents answering "yes"<br>(weighted %) |  |
| --- | --- | --- |
|  | UK (n=1768) | HK (n=1663) |
| **Contact** |  |  |
| Face-to-face conversation (no physical contact) with someone who has coronavirus but no symptoms | 1234 (69.2) | 1564 (94.0) |
| Face-to-face conversation (no physical contact) with someone who has coronavirus with symptoms | 1398 (78.7) | 1616 (96.7) |
| Physical contact with someone who has coronavirus but no symptoms | 1580 (89.0) | 1635 (98.1) |
| Physical contact with someone with coronavirus who has symptoms | 1657 (93.5) | 1644 (98.5) |
| **Transmission mode** |  |  |
| Droplets | - | 1649 (99.2) |
| Aerosol when infected people cough or sneeze | - | 1478 (91.3) |
| Being in close contact (i.e. within 2 meters) to someone who has coronavirus when they cough or sneeze | 1604 (90.4) | - |
| Being further away (i.e. further than 2 meters away) to someone who has coronavirus, when they cough or sneeze | 615 (34.8) | - |
| **Others** |  |  |
| Contact with virus-contaminated environment | 1411 (79.5) | 1594 (96.2) |
| Consumption of wild animal meat | 199 (11.3) | 1546 (93.3) |
| Visiting wet market | 374 (21.5) | 1342 (80.9) |
| Consumption of seafood imported from specific regions <sup>a</sup> | 258 (14.8) | 1199 (70.7) |
| Consumption/use of products imported from specific regions <sup>a</sup> | 209 (12.1) | 1101 (66.5) |

<sup>a</sup> Specific regions refer to Wuhan (HK)/ China (UK).

### Adoption of social distancing measures

There were variations in the weighted proportion of HK and the UK respondents adopting precautionary measures against COVID-19 (**Figure 1**). HK respondents reported higher levels of adoption across all social distancing and personal hygiene measures. In particular, 98.8% of HK respondents reported wearing a face mask compared to 3.1% among the UK respondents. General measures were adopted by 68.1-87.3 % and 37.8-59.0 % of respondents in HK and the UK, respectively. Contact measures were adopted by 83.9-93.7% and 33.7-50.1 % of respondents in HK and the UK, respectively. Work measure was reported by 32.4% and 22.5% of respondents in HK and the UK, respectively.

**Figure 1.**
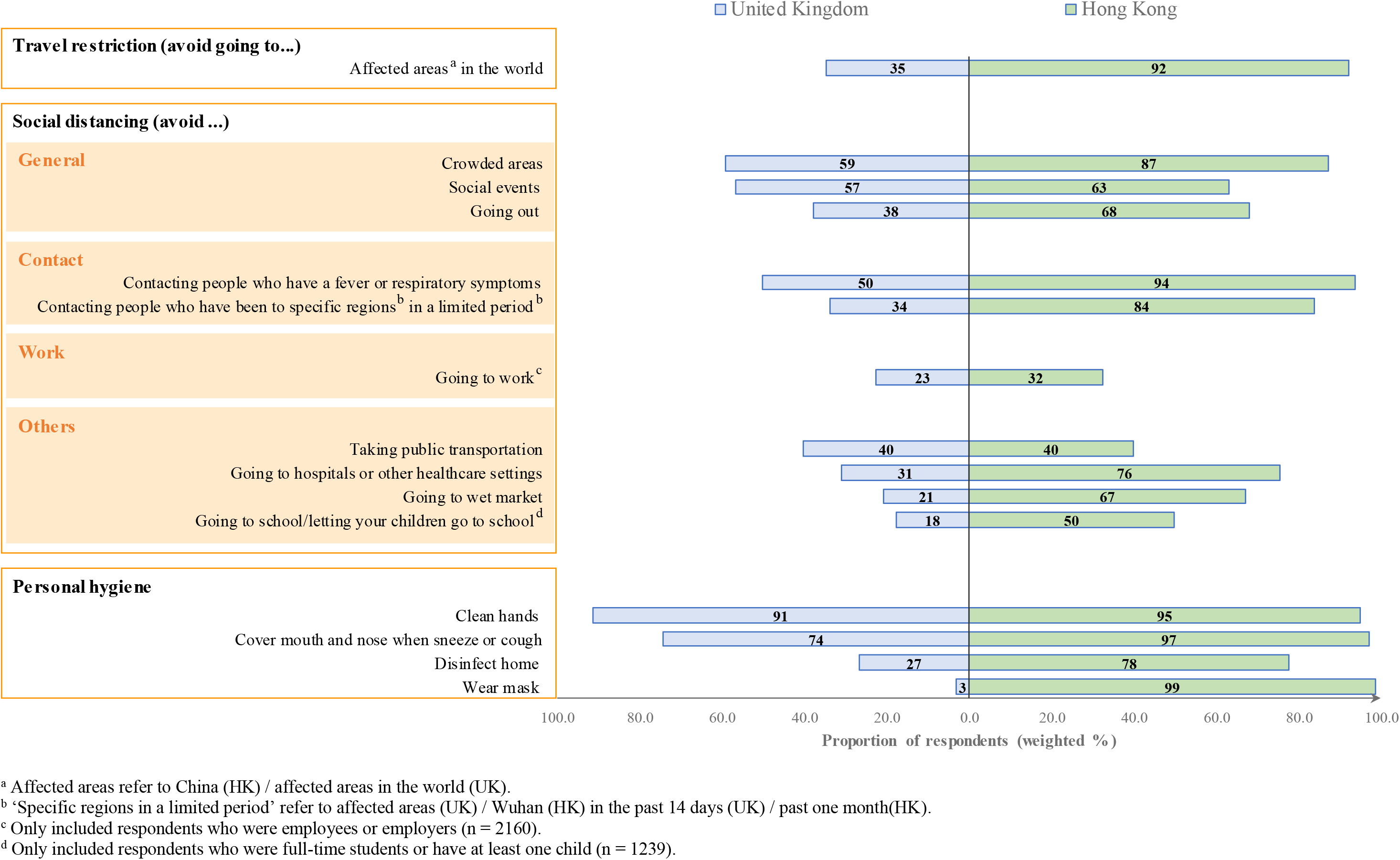
Adoption of precautionary measures against COVID-19

Sociodemographic factors were associated with the three social distancing measures (**Table S2; Table 3**). The UK respondents were significantly less likely than their HK counterparts to adopt social distancing measures (**Table S2**, aOR: 0.08-0.53, *P*<0.001; **Table 3**, aOR:0.08-0.70, *P*<0.001). When adjusting for regional differences, general measures were less likely to be adopted by males (aOR:0.82; 95% CI:0.71-0.95) but more likely to be adopted by the unemployed (aOR:1.65; 95% CI: 1.30-2.09) or retired (aOR:1.92; 95% CI: 1.43-2.59). Contact measures were less likely to be adopted by males (aOR:0.74; 95% CI:0.63-0.88) but more likely to be adopted by those retired (aOR:1.40; 95% CI: 1.03-1.91). Finally, work measure was less likely to be adopted by those aged 55 years or above (aOR:0.60; 95% CI: 0.39-0.93).

**Table 3.** Factors associated with the adoption of different types of social distancing

| Factors | Types of social distancing |  |  |  |  |  |
| --- | --- | --- | --- | --- | --- | --- |
|  | General <sup>a</sup> (n=3431)<br>(Model 1) |  | Contact <sup>b</sup> (n=3431)<br>(Model 2) |  | Work <sup>c</sup> (n=2160)<br>(Model 3) |  |
|  | aOR (95% CI) | P-value | aOR (95% CI) | P-value | aOR (95% CI) | P-value |
| **Age** |  |  |  |  |  |  |
| 18-24 | - | - | - | - | - | - |
| 25-34 | 1.54 (1.16, 2.05) | 0.003 | 0.96 (0.68, 1.35) | 0.81 | 1.00 (0.72, 1.39) | 0.99 |
| 35-44 | 1.25 (0.93, 1.68) | 0.13 | 0.74 (0.53, 1.05) | 0.09 | 0.95 (0.68, 1.33) | 0.77 |
| 45-54 | 1.30 (0.95, 1.79) | 0.10 | 0.67 (0.47, 0.97) | 0.03 | 0.72 (0.49, 1.05) | 0.09 |
| 55+ | 0.99 (0.70, 1.41) | 0.97 | 0.81 (0.55, 1.19) | 0.28 | 0.60 (0.39, 0.93) | 0.02 |
| **Sex** |  |  |  |  |  |  |
| Female | - | - | - | - | - | - |
| Male | 0.82 (0.71, 0.95) | 0.01 | 0.74 (0.63, 0.88) | <0.001 | 0.95 (0.78, 1.16) | 0.62 |
| **Education attainment <sup>d</sup>** |  |  |  |  |  |  |
| No formal qualification/<br>lower secondary or below | - | - | - | - | - | - |
| Secondary level qualification/<br>higher secondary | 0.99 (0.68, 1.44) | 0.94 | 0.96 (0.64, 1.44) | 0.85 | 1.04 (0.46, 2.34) | 0.92 |
| Post-secondary but below degree | 1.06 (0.72, 1.55) | 0.78 | 1.12 (0.74, 1.71) | 0.58 | 1.09 (0.48, 2.47) | 0.83 |
| Degree or above | 1.27 (0.87, 1.83) | 0.21 | 0.98 (0.66, 1.47) | 0.94 | 1.94 (0.88, 4.28) | 0.10 |
| **Employment status** |  |  |  |  |  |  |
| Employed | - | - | - | - | - | - |
| Full-time student | 1.35 (0.98, 1.85) | 0.07 | 1.08 (0.73, 1.59) | 0.70 | - | - |
| Unemployed | 1.65 (1.30, 2.09) | <0.001 | 1.20 (0.91, 1.58) | 0.20 | - | - |
| Retired | 1.92 (1.43, 2.59) | <0.001 | 1.40 (1.03, 1.91) | 0.03 | - | - |
| **Region** |  |  |  |  |  |  |
| HK | - | - | - | - | - | - |
| UK | 0.35 (0.30, 0.41) | <0.001 | 0.08 (0.07, 0.10) | <0.001 | 0.70 (0.57, 0.87) | <0.001 |

<sup>a</sup> "General" refers to avoiding going to: (i) crowded areas; (ii) social events; and (iii) going out.
<sup>b</sup> "Contact " refers to avoiding contacting individuals who (i) had a fever or respiratory symptoms; and (ii) had been to affected areas (UK) / Wuhan (HK) in the past 14 days (UK) / past month (HK).
<sup>c</sup> "Work" refers to avoiding going to work.

The impact of perceived severity of infection and perceived ease of transmission on the adoption of social distancing behaviours vary by region (**Table 4**). In HK, they had no impact, whereas in the UK, those who perceived COVID-19 infection as serious were more likely to adopt all social distancing measures (aOR:1.58-3.01), and those who perceived transmission of SARS-CoV-2 as easy were more prone to adopt both general social distancing measures (Model 4.2, aOR:2.00, 95% CI:1.57-2.55) and contact measures (Model 5.2, aOR:1.80, 95% CI:1.41-2.30). On the other hand, the impact of anxiety on the adoption of social distancing behaviours did not significantly differ by region (**Table 4**). Those with borderline abnormal (HK, aOR: 1.26-1.62; UK, aOR: 1.36-1.48) or abnormal HADS-A (HK, aOR:1.82-2.09; UK, aOR:1.40-2.40) scores were more likely to adopt all three types of social distancing measures compared to those with normal anxiety levels.

**Table 4.** Region-specific effect and the effect modification (by regions) of selected variables on the adoption of social distancing

| Region | Selected variables | Types of social distancing |  |  |  |  |  |
| --- | --- | --- | --- | --- | --- | --- | --- |
|  |  | General <sup>a</sup> (n=3431) |  | Contact <sup>b</sup> (n=3431) |  | Work <sup>c</sup> (n=2160) |  |
|  |  | aOR (95% CI) | p-value | aOR (95% CI) | p-value | aOR (95% CI) | p-value |
|  | **Perceived severity <sup>d</sup>** | **(Model 4.1) <sup>e</sup>** |  | **(Model 5.1) <sup>e</sup>** |  | **(Model 6.1) <sup>e</sup>** |  |
| HK | Not serious | - | - | - | - | - | - |
|  | Serious | 0.93 (0.50, 1.74) | 0.82 | 1.71 (0.85, 3.47) | 0.13 | 0.63 (0.30, 1.30) | 0.21 |
| UK | Not serious | - | - | - | - | - | - |
|  | Serious | 3.01 (2.35, 3.86) | <0.001 | 1.90 (1.48, 2.43) | <0.001 | 1.58 (1.06, 2.37) | 0.03 |
|  | Effect modification <sup>f</sup> | 3.24 (1.65, 6.35) | <0.001 | 1.11 (0.52, 2.34) | 0.79 | 2.52 (1.09, 5.80) | 0.03 |
|  | **Perceived ease of transmission <sup>g</sup>** | **(Model 4.2) <sup>e</sup>** |  | **(Model 5.2) <sup>e</sup>** |  | **(Model 6.2) <sup>e</sup>** |  |
| HK | Difficult | - | - | - | - | - | - |
|  | Easy | 1.15 (0.77, 1.74) | 0.50 | 1.00 (0.58, 1.71) | 0.99 | 0.61 (0.37, 1.03) | 0.07 |
| UK | Difficult | - | - | - | - | - | - |
|  | Easy | 2.00 (1.57, 2.55) | <0.001 | 1.80 (1.41, 2.30) | <0.001 | 1.34 (0.96, 1.87) | 0.09 |
|  | Effect modification <sup>f</sup> | 1.73 (1.07, 2.79) | 0.02 | 1.81 (1.00, 3.28) | 0.05 | 2.18 (1.18, 4.04) | 0.01 |
|  | **Anxiety level** | **(Model 4.3) <sup>e</sup>** |  | **(Model 5.3) <sup>e</sup>** |  | **(Model 6.3) <sup>e</sup>** |  |
| HK | Normal | - | - | - | - | - | - |
|  | Borderline abnormal | 1.62 (1.27, 2.06) | <0.001 | 1.26 (0.93, 1.70) | 0.14 | 1.51 (1.10, 2.06) | 0.01 |
|  | Abnormal | 2.09 (1.64, 2.66) | <0.001 | 1.85 (1.34, 2.56) | <0.001 | 1.82 (1.34, 2.48) | <0.001 |
| UK | Normal | - | - | - | - | - | - |
|  | Borderline abnormal | 1.48 (1.11, 1.96) | 0.007 | 1.36 (1.02, 1.80) | 0.03 | 1.37 (0.92, 2.02) | 0.12 |
|  | Abnormal | 2.40 (1.87, 3.09) | <0.001 | 1.76 (1.37, 2.27) | <0.001 | 1.40 (0.99, 1.98) | 0.06 |
|  | Effect modification (borderline abnormal) <sup>f</sup> | 0.91 (0.63, 1.33) | 0.64 | 1.08 (0.71, 1.63) | 0.71 | 0.91 (0.55, 1.49) | 0.70 |
|  | Effect modification (abnormal) <sup>f</sup> | 1.15 (0.82, 1.62) | 0.42 | 0.95 (0.63, 1.42) | 0.80 | 0.77 (0.48, 1.21) | 0.26 |

<sup>a</sup> "General" refers to avoiding going to: (i) crowded areas; (ii) social events; and (iii) going out.
<sup>b</sup> "Contact" refers to avoiding contacting individuals who (i) had a fever or respiratory symptoms; and (ii) had been to affected areas (UK) / Wuhan (HK) in the past 14 days (UK) / past one month (HK).
<sup>c</sup> "Work" refers to avoiding going to work.
<sup>d</sup> For perceived severity, "serious" refers to levels 1-2; whereas "not serious" refers to levels 3-5. (Refer to footnote b in Table 1)
<sup>e</sup> The models have been adjusted for all covariates in Table 4.
<sup>f</sup> It measures the "difference of the effect being considered" due to "difference in region", and its value is the ratio of the two region-specific effects.
<sup>g</sup> For perceived ease of transmission, 'ease' refers to that the virus can be transmitted by having face-to face conversation with someone who has coronavirus but no symptoms; otherwise, the perceived ease of transmission is 'difficult'.

## DISCUSSION

### Principal results

This study compared the initial public perceptions and preventative behaviours during the COVID-19 pandemic across HK and the UK. The adoption rate of social-distancing measures was higher in HK than in the UK. Risk perception and knowledge of COVID-19 were consistently and significantly higher in HK, but the UK respondents were more likely to alter behaviours to align with these two metrics: where transmission was considered ‘easy’, and perceived severity was considered ‘severe’, they were more likely to adopt preventive behaviours. Anxiety was a consistent driver of behaviour change irrespective of context: those more anxious were more likely to adopt preventative actions. Such behaviour is consistent with the wider literature surrounding the adoption of precautionary measures **[6, 7]**, and in particular, provides further evidence that anxiety drives protective behaviours, such as handwashing **[8]**, an effective intervention against the transmission of respiratory diseases **[22]**.

### Implications

This study has three implications. First, health officials should account for region-specific baseline levels of risk perception and knowledge when designing and promoting mitigation strategies. The evidence presented by this study demonstrates that regional context is important in terms of both how people understand risk, and how risk drives behaviour. While the significant social, historical and cultural heterogeneity between HK and the UK likely contributes to the results observed in this study, the importance of additional intrinsic factors, such as population sensitisation via past infectious disease outbreaks, and state-led health promotion campaigns, should not be underestimated. In studies elsewhere, public perceptions of these factors have been significant drivers of adoption of preventative behaviours during previous epidemics **[23]**, while conceptions of personal risk have been connected to individuals’ understanding of local disease prevalence and severity [**24–26**]. Therefore, assessment of the baseline population knowledge, attitudes and practices (KAP) and subsequent continual monitoring throughout the pandemic are essential to any effective context-specific pandemic preparedness plan.

Second, risk communication should build upon baseline KAP outcomes and develop trust across suitable media channels. Significant contextual heterogeneity in the public reliance on information sources provides additional insight here. HK reported greater reliance on social media and far less trust in official websites, suggesting that official messaging in HK unlikely drove individual behaviour change; by contrast, the UK results suggested that although the UK government held an effective platform to influence public health behaviour, government health messaging was insufficient to attain similar HK baseline knowledge levels, particularly in the absence of prior population sensitisation to infectious disease outbreaks. Therefore, there is a pressing need to tailor communication approaches, likely on a graduated scale, but as a minimum in a binary fashion to accommodate both “naive” and “experienced” populations.

Third, the comparative snapshots of initial community responses captured by this study demonstrate the diversity in approach and pandemic response during the early phases. Across many contexts, national lockdowns became commonplace as the true magnitude of transmission became apparent; but the associated indirect costs render blanket strategies untenable in the medium term. As national lockdowns are lifted, countries worldwide face the challenge of resurging cases, and must consider nuanced approaches to prevent additional harm. Driven by anxiety, high perceived severity and knowledge, HK conducted widespread preventative measures early, and en masse. Together with early government actions **[27]**, the strategies adopted by the HK community were successful during the initial phase of the pandemic. Considering this, and that national populations are now highly sensitised to COVID-19 transmission, tailored public health messaging, early regional containment, and increased health capacity should ensure more effective public health responses with less indirect impact on national economies.

### Study strengths and limitations

This study is one of few that compared community responses during a pandemic using similar metrics while adjusting for disease progression. It has two limitations though. First, both samples varied significantly across the demographic spectrum, and while responses were weighted, such convenience sampling is unlikely to be representative of the wider populations. Second, the survey period in HK was three weeks, compared to two days in the UK. Therefore, while the timing of the two surveys relative to each national epidemic was comparable, there may be temporal differences between survey duration, which might lead to some sampling bias, especially during the initial phase of the pandemic when there is much uncertainty about the disease.

### Conclusions

To conclude, this study compared the initial community responses towards COVID-19 in HK and the UK. In line with the high baseline level of risk perception and knowledge, and historical exposure to respiratory disease outbreaks, the adoption of preventive measures was higher in HK. However, the UK sample demonstrated that the adoption rate of preventive behaviours could be improved by heightened risk perception and knowledge, best driven by improved public health campaigns. Together, these results suggest that health officials should ascertain and consider baseline levels of risk perception and knowledge, as well as prior sensitisation to infectious disease outbreaks, during the development of mitigation strategies. Risk communication should be done through suitable media channels - and trust should be maintained - while early intervention remains the cornerstone of effective outbreak response.

## Data Availability

Please contact the corresponding authors about the research data which generates the study results.

## CONTRIBUTION

Bowman LR (BLR), Kwok KO (KKO), Ward H (WH), Atchison C (AC) and Wong SYS (WSYS) conceived the study; KKO, Wei WI (WWI) and AC collected the data; KKO, Yi YY (YYY) and WWI analysed the data; BLR, KKO, Redd RE (RRE), WH, WWI, AC and WSYS interpreted the data; BLR wrote the first draft of the manuscript; KKO, RRE, YYY, WH, WWI, AC, and WSYS edited the manuscript.

## ACKNOWLEDGEMENTS

The study was supported by Imperial NIHR Research Capability Funding and the internal funding of The Chinese University of Hong Kong.

## CONFLICTS OF INTEREST

The authors declare that they have no known competing financial interests or personal relationships that could have appeared to influence the work reported in this paper.

**ABBREVIATIONS**
HK: Hong Kong
UK: United Kingdom
COVID-19: coronavirus disease 2019
SARS: severe acute respiratory syndrome
HASD-A: Hospital, Anxiety and Depression scale - Anxiety
aOR: adjusted odds ratio
CI: confidence interval
KAP: knowledge, attitudes and practices

## REFERENCES

1. World Health Organization. Rolling updates on coronavirus disease (COVID-19) (Updated 31 July 2020) 2020 [Available from: https://www.who.int/emergencies/diseases/novel-coronavirus-2019/events-as-they-happen.

2. Hong Kong Centre for Health Protection. Countries/areas with reported cases of Coronavirus Disease-2019 (COVID-19) (Last updated on August 2, 2020, 11 am) 2020 [Available from: https://www.chp.gov.hk/files/pdf/statistics_of_the_cases_novel_coronavirus_infection_en.pdf

3. Kwok KO, Lai FYL, Wei VWI, Tsoi MTF, Wong SYS, Tang JWT. Comparing the impact of various interventions to control the spread of COVID-19 in twelve countries. J Hosp Infect. 2020. PMID: 32619456

4. Prime Minister’s Office of The governemnt of the United Kingdom. Prime Minister’s statement on coronavirus (COVID-19): 16 March 2020 2020 [Available from: https://www.gov.uk/government/speeches/pm-statement-on-coronavirus-16-march-2020.

5. Kwok KO, Li KK, Chan HHH, Yi YY, Tang A, Wei WI, et al. Community Responses during Early Phase of COVID-19 Epidemic, Hong Kong. Emerg Infect Dis. 2020;26(7):1575–9. PMID: 32298227

6. Leung GM, Ho LM, Chan SK, Ho SY, Bacon-Shone J, Choy RY, et al. Longitudinal assessment of community psychobehavioral responses during and after the 2003 outbreak of severe acute respiratory syndrome in Hong Kong. Clin Infect Dis. 2005;40(12):1713–20. PMID: 15909256

7. Bish A, Michie S. Demographic and attitudinal determinants of protective behaviours during a pandemic: a review. Br J Health Psychol. 2010;15(Pt 4):797–824. PMID: 20109274

8. Jones JH, Salathe M. Early assessment of anxiety and behavioral response to novel swine-origin influenza A(H1N1). PLoS One. 2009;4(12):e8032. PMID: 19997505

9. Bults M, Beaujean DJ, de Zwart O, Kok G, van Empelen P, van Steenbergen JE, et al. Perceived risk, anxiety, and behavioural responses of the general public during the early phase of the Influenza A (H1N1) pandemic in the Netherlands: results of three consecutive online surveys. BMC Public Health. 2011;11:2. PMID: 21199571

10. Lohiniva AL, Sane J, Sibenberg K, Puumalainen T, Salminen M. Understanding coronavirus disease (COVID-19) risk perceptions among the public to enhance risk communication efforts: a practical approach for outbreaks, Finland, February 2020. Euro Surveill. 2020;25(13). PMID: 32265008

11. Gesser-Edelsburg A, Cohen R, Hijazi R, Abed Elhadi Shahbari N. Analysis of Public Perception of the Israeli Government’s Early Emergency Instructions Regarding COVID-19: Online Survey Study. J Med Internet Res. 2020;22(5):e19370. PMID: 32392172

12. Motta Zanin G, Gentile E, Parisi A, Spasiano D. A Preliminary Evaluation of the Public Risk Perception Related to the COVID-19 Health Emergency in Italy. Int J Environ Res Public Health. 2020;17(9). PMID: 32349253

13. Olapegba PO, Ayandele O. Survey data of COVID-19-related Knowledge, Risk Perceptions and Precautionary Behavior among Nigerians. Data Brief. 2020:105685. PMID: 32391411

14. McFadden SM, Malik AA, Aguolu OG, Willebrand KS, Omer SB. Perceptions of the adult US population regarding the novel coronavirus outbreak. PLoS One. 2020;15(4):e0231808. PMID: 32302370

15. Huynh TLD. Data for understanding the risk perception of COVID-19 from Vietnamese sample. Data Brief. 2020;30:105530. PMID: 32322641

16. Atchison CJ, Bowman L, Vrinten C, Redd R, Pristera P, Eaton JW, et al. Perceptions and behavioural responses of the general public during the COVID-19 pandemic: A cross-sectional survey of UK Adults. 2020.

17. Hong Kong Centre for Health Protection. Latest situation of cases of COVID-19 (as of 2 August 2020) 2020 [Available from: https://www.chp.gov.hk/files/pdf/local_situation_covid19_en.pdf.

18. Department of Health and Social Care and Public Health England. Coronavirus cases in the UK: daily updated statistics 2020 [Available from: https://www.gov.uk/guidance/coronavirus-covid-19-information-for-the-public.

19. Snaith RP. The Hospital Anxiety And Depression Scale. Health Qual Life Outcomes. 2003;1:29. PMID: 12914662

20. Office for National Statistics. 2011 Census 2011 [Available from: https://www.ons.gov.uk/census/2011census.

21. The Census and Statistics Department. By-census Results 2016 [Available from: https://www.bycensus2016.gov.hk/en/.

22. Rabie T, Curtis V. Handwashing and risk of respiratory infections: a quantitative systematic review. Trop Med Int Health. 2006;11(3):258–67. PMID: 16553905

23. SteelFisher GK, Blendon RJ, Ward JR, Rapoport R, Kahn EB, Kohl KS. Public response to the 2009 influenza A H1N1 pandemic: a polling study in five countries. Lancet Infect Dis. 2012;12(11):845–50. PMID: 23041199

24. Funk S, Salathe M, Jansen VA. Modelling the influence of human behaviour on the spread of infectious diseases: a review. J R Soc Interface. 2010;7(50):1247–56. PMID: 20504800

25. de Zwart O, Veldhuijzen IK, Elam G, Aro AR, Abraham T, Bishop GD, et al. Perceived threat, risk perception, and efficacy beliefs related to SARS and other (emerging) infectious diseases: results of an international survey. Int J Behav Med. 2009;16(1):30–40. PMID: 19125335

26. Bults M, Beaujean DJ, Richardus JH, Voeten HA. Perceptions and behavioral responses of the general public during the 2009 influenza A (H1N1) pandemic: a systematic review. Disaster Med Public Health Prep. 2015;9(2):207–19. PMID: 25882127

27. Wong SYS, Kwok KO, Chan FKL. What can countries learn from Hong Kong’s response to the COVID-19 pandemic? CMAJ. 2020;192(19):E511–E5. PMID: 32332040

